# Clinical Characteristics and Prognostic Factors for ICU Admission of Patients with Covid-19 Using Machine Learning and Natural Language Processing

**DOI:** 10.1101/2020.05.22.20109959

**Authors:** Jose L. Izquierdo, Julio Ancochea, Savana COVID-19 Research Group, Joan B. Soriano

## Abstract

There remain many unknowns regarding the onset and clinical course of the ongoing COVID-19 pandemic. We used a combination of classic epidemiological methods, natural language processing (NLP), and machine learning (for predictive modeling), to analyse the electronic health records (EHRs) of patients with COVID-19.

We explored the unstructured free text in the EHRs within the SESCAM Healthcare Network (Castilla La-Mancha, Spain) from the entire population with available EHRs (1,364,924 patients) from January 1^st^ to March 29^th^, 2020. We extracted related clinical information upon diagnosis, progression and outcome for all COVID-19 cases, focusing in those requiring ICU admission.

A total of 10,504 patients with a clinical or PCR-confirmed diagnosis of COVID-19 were identified, 52.5% males, with age of 58.2±19.7 years. Upon admission, the most common symptoms were cough, fever, and dyspnoea, but all in less than half of cases. Overall, 6% of hospitalized patients required ICU admission. Using a machine-learning, data-driven algorithm we identified that a combination of age, fever, and tachypnoea was the most parsimonious predictor of ICU admission: those younger than 56 years, without tachypnoea, and temperature <39°C, (or >39°C without respiratory crackles), were free of ICU admission. On the contrary, COVID-19 patients aged 40 to 79 years were likely to be admitted to the ICU if they had tachypnoea and delayed their visit to the ER after being seen in primary care.

Our results show that a combination of easily obtainable clinical variables (age, fever, and tachypnoea with/without respiratory crackles) predicts which COVID-19 patients require ICU admission.

## INTRODUCTION

The unprecedented, global spread of the severe acute respiratory syndrome coronavirus 2 (SARS-CoV-2) that causes coronavirus disease 2019 (COVID-19) requires innovative approaches that deliver immediate, real-time results[1, 2]. To date, big data technologies have only been used to estimate SARS-CoV-2 transmission[3], and to indirectly estimate COVID-19 cases in China by using social media[4]. However, there remain many unknowns regarding the onset and temporal distribution of the ongoing COVID-19 pandemic. Similarly, both the individual and population burden of COVID-19 are just beginning to be unravelled. To the best of our knowledge, such tools[5-7] have not been used to explore the clinical characteristics and prognostic factors of COVID-19[8].

Considering the unprecedented spread and severity of the ongoing COVID-19 outbreak, focus has been set on hospital’s unmet need, and in particular ICU requirements[8, 9]. Indeed, health systems have been/are near collapse and independent modelling efforts have aimed to forecast a number of epidemiological estimators, including ICU use [10-12].

Previously, our team reported that the combination of big data analytics and machine learning techniques helped to better determine quality of diagnosis and treatment of chronic obstructive pulmonary disease (COPD) via an analysis of hospital electronic health records (EHRs) using natural language processing (NLP) and validated algorithms[13, 14].

By means of The BigCOVIData study, we aimed to better determine the real-world epidemiology of COVID-19 infection in a well-defined population. Using a combination of classic epidemiological methods[15], NLP, and machine learning (for predictive modeling), we analysed the clinical information contained in the EHRs of patients with COVID-19 to advance our understanding of the disease and its associated outcomes, most notably ICU admission.

## METHODS

The BigCOVIData study was conducted in compliance with legal and regulatory requirements and followed generally accepted research practices described in the ICH Guideline for Good Clinical Practice, the Helsinki Declaration in its latest edition, Good Pharmacoepidemiology Practices, and applicable local regulations. This study was classified as a ‘non-post-authorization study’ (EPA) by the Spanish Agency of Medicines and Health Products (AEMPS), and it was approved by the Research Ethics Committee at the University Hospital of Guadalajara (Spain). We have followed and endorsed the STrengthening the Reporting of OBservational studies in Epidemiology (STROBE) guidance for reporting observational research[16].

### Study design and data source

This was a multicenter, non-interventional, retrospective study using data captured in the EHRs of the participating hospitals within the SESCAM Healthcare Network in Castilla-La Mancha, Spain (**Figure 1**). Data captured in the EHRs was collected from all available departments, including inpatient hospital, outpatient hospital, and ER, for virtually all types of provided services in each participating hospital. The study period was January 1, 2020 – March 29, 2020.

**Figure 1.**
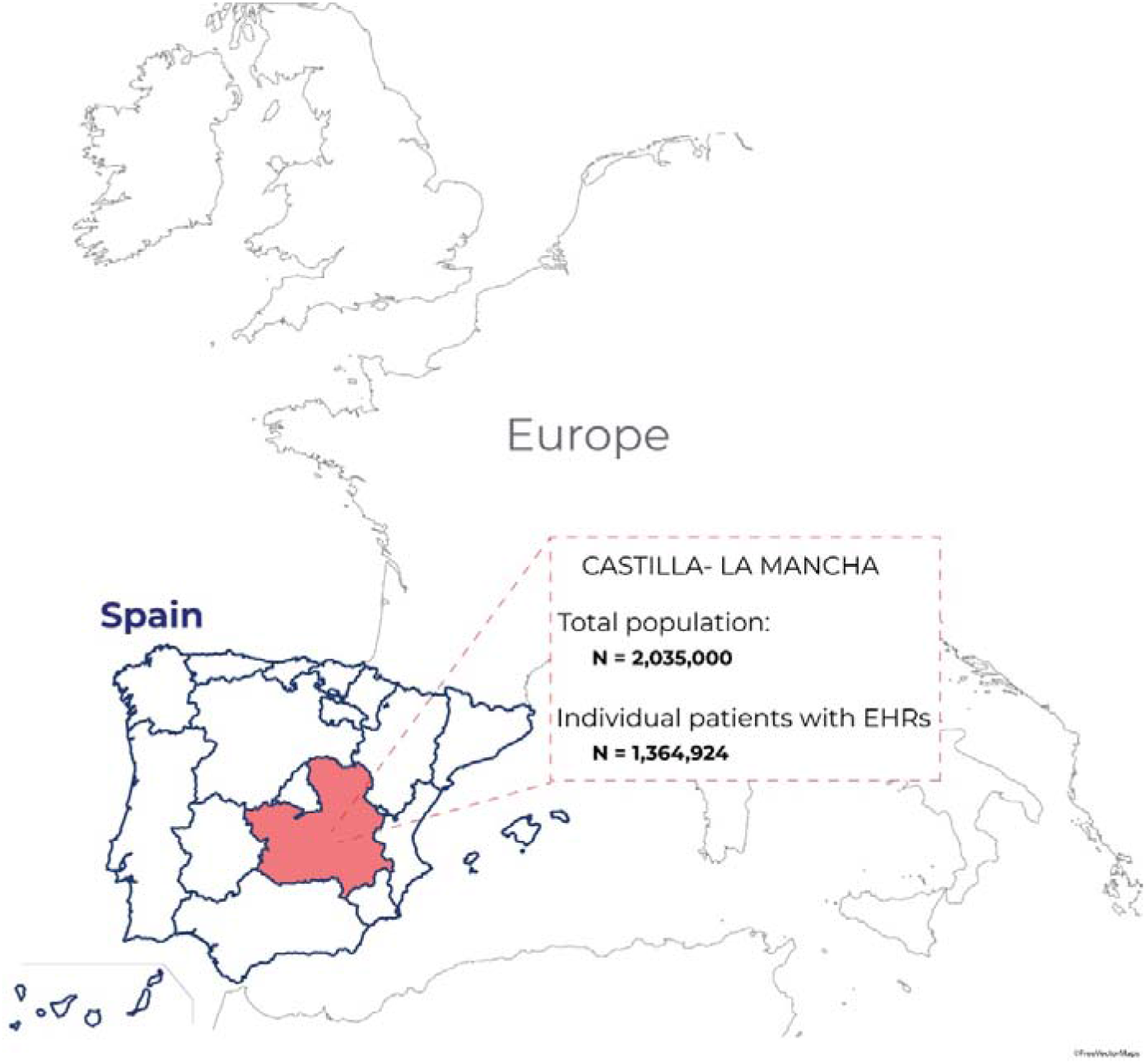
Map of Castilla-La Mancha. **Footnote**: Map of Castilla-La Mancha (red) within the Spanish (blue line) and European territories. From a source general population of 2,035,000 inhabitants, we collected and analyzed the clinical information in the EHRs of 1,364,924 patients within the SESCAM Healthcare Network.

The study database was fully anonymized in a structured format and contained no personal information from patients. Likewise, personal information was not accessed during either the application of automated and algorithmic methods (i.e., NLP) or during the conversion of unstructured data into the structured database. Importantly, given that clinical information was handled in an aggregate, anonymized, and irreversibly dissociated manner, patient consent regulations do not apply to the present study

### Study sample

The study sample included all patients in the source population diagnosed with COVID-19. Patients were identified on the basis of clinical diagnosis (i.e., COVID-19 cases determined by observed symptomatology, imaging (mostly chest X-ray) and laboratory results, as captured in the unstructured, free-text information in the EHRs) and/or microbiological test results (i.e., COVID-19 cases confirmed by RT-PCR or similar available tests). Our decision to consider both PCR- and clinically confirmed cases is justified by the limited availability of routinely administered RT-PCR tests in the region during the study period and supported by recent discussions on the far-from-optimal sensitivity of RT-PCR for COVID19 (i.e., a single negative result from a single specimen cannot exclude the disease in suspected cases)[17, 18]. Indeed, recent reports highlight the clinical validity and relatively high sensitivity of symptom- and imaging-based identification of COVID-19 patients, especially in early stages of the disease[17, 19, 20].

### EHRead®

To meet the study objectives, we used EHRead®[21], a technology developed by SAVANA that applies NLP, machine learning, and deep learning to analyse the unstructured free-text information written in millions of de-identified EHRs. This technology enables the extraction of information from all types of EHRs and the subsequent normalization of extracted clinical entities to a unique terminology. This process allows for further analysis of descriptive or predictive nature. Originally based on SNOMED CT terminology, our unique body of terminology comprises more than 400,000 medical concepts, acronyms, and laboratory parameters aggregated over the course of five years of free-text mining, targeting the most common diseases (e.g. respiratory diseases, cardiovascular diseases, and diabetes, among others).

Using a combination of regular expression (regex) rules and machine learning models, the terminology entities are detected in the unstructured text and later classified based on sections typically contained in the EHRs, hospital services, and other clinical specifications. Importantly, each detected term is described in terms of negative, speculative, or affirmative clinical statements; this is achieved by using deep learning CNN classification methods that rely on word embeddings and context information (for a similar methodological approach, see [22]). Limitations in a case by case detection are also overcome with a similar approach to ensure that the detected concepts are used within the appropriate context for the descriptive and predictive analysis.

For particular cases where extra specifications are required (i.e., to differentiate COVID cases from other mentions of the term related to fear of the disease or to potential contact), the detection output was manually reviewed in more than 5000 reports to avoid any possible ambiguity associated with free-text reporting. All NLP deep learning models used in this study were validated using the standard training/validation/testing approach; we used a 75/12/13 split ratio in the available annotated data (between 2,000 and 3,000 records, depending on the model) to ensure efficient generalization on unseen cases. For all developed models, we obtained F-scores greater than 0.89.

### Data Analyses

All categorical variables (e.g., comorbidities, symptoms) are shown in frequency tables, whereas continuous variables (e.g., age) are described via summary tables that include the mean, standard deviation, median, minimum, maximum, and quartiles of each variable. The number of missing data points for each variable is provided, if any. To test for possible statistically significant differences in the distribution of categorical variables between study groups (i.e., male vs. female, ICU admission vs. No ICU admission), we used Yates-corrected chi^2^ tests. For continuous variables, mean differences were tested using t-tests. Given our general population approach, and our larger than usual sample size, we were interested in exploring sex-related differences in COVID-19 patients, so most results are stratified by sex[23]. All statistical inferences were performed at the 5% significance level using 2-sided tests or 2-sided CIs.

### Predictive model

We developed a decision tree to classify COVID-19 patients according to their risk of being admitted to the ICU. The two types of patients or *classes* considered in the model were therefore “admitted to the ICU” and “not admitted to the ICU”. The model maps the characteristics of patients (the *variables)* to their class in the shape of a tree. From a clinical perspective, this model contemplates all patient variables upon admission, meaning that its predictive value is so from symptom debut until outcome. The tree is composed of nodes that branch to subsequent children nodes depending on the patient’s variables. The tree is built in such a way that each branch separates the two classes as much as possible. This separation is measured as *Shannon entropy*, where a node with an entropy of zero means that the classification is perfect (either all or none of the patients were admitted to the ICU) and an entropy of one is the worst possible mix (50%/50%)[24].

#### Model training and validation

The model was developed and tested on the available data from hospitalized patients that had either been admitted to the ICU or not; the latter were either discharged from the hospital or died in the course of the disease. This amounted to a total of 900 patients. Our algorithm was validated in a split of our COVID-19 sample, in a 70% training set and a 30% validation set. This means that the model was trained with 630 patients (582 who did not require intensive care, vs 48 who did) and validated over the remaining 270 patients. Because the two classes were unbalanced (far fewer patients require ICU), we used the standard technique of oversampling the lower class to guarantee a balance of accuracy and recall (in other words, the tradeoff between false positives vs. false negatives). Further, we sought to replicate the results from this validation in *a posteriori* sensitivity analysis, as per recent recommendations for predictive modeling in COVID-19[25] and TRIPOD guidance[26]. For this second validation, we trained the model with data from the provinces of Ciudad Real and Guadalajara (38% of the study sample from Castilla La-Mancha), and used an independent set with combined data set from the other three provinces, namely Toledo, Cuenca, and Albacete for validation.

## RESULTS

From a source general population of 2,035,000 inhabitants, we used NLP and machine learning to analyse the clinical information contained in the EHRs of 1,364,924 anonymous patients (**Figure 1**). Among these, we identified a total of 10,504 patients diagnosed with COVID-19 (**Figure 2**). The flowchart of participation in the study up to hospital admission, ICU admission, or discharge is presented in **Figure 2**.

**Figure 2.**
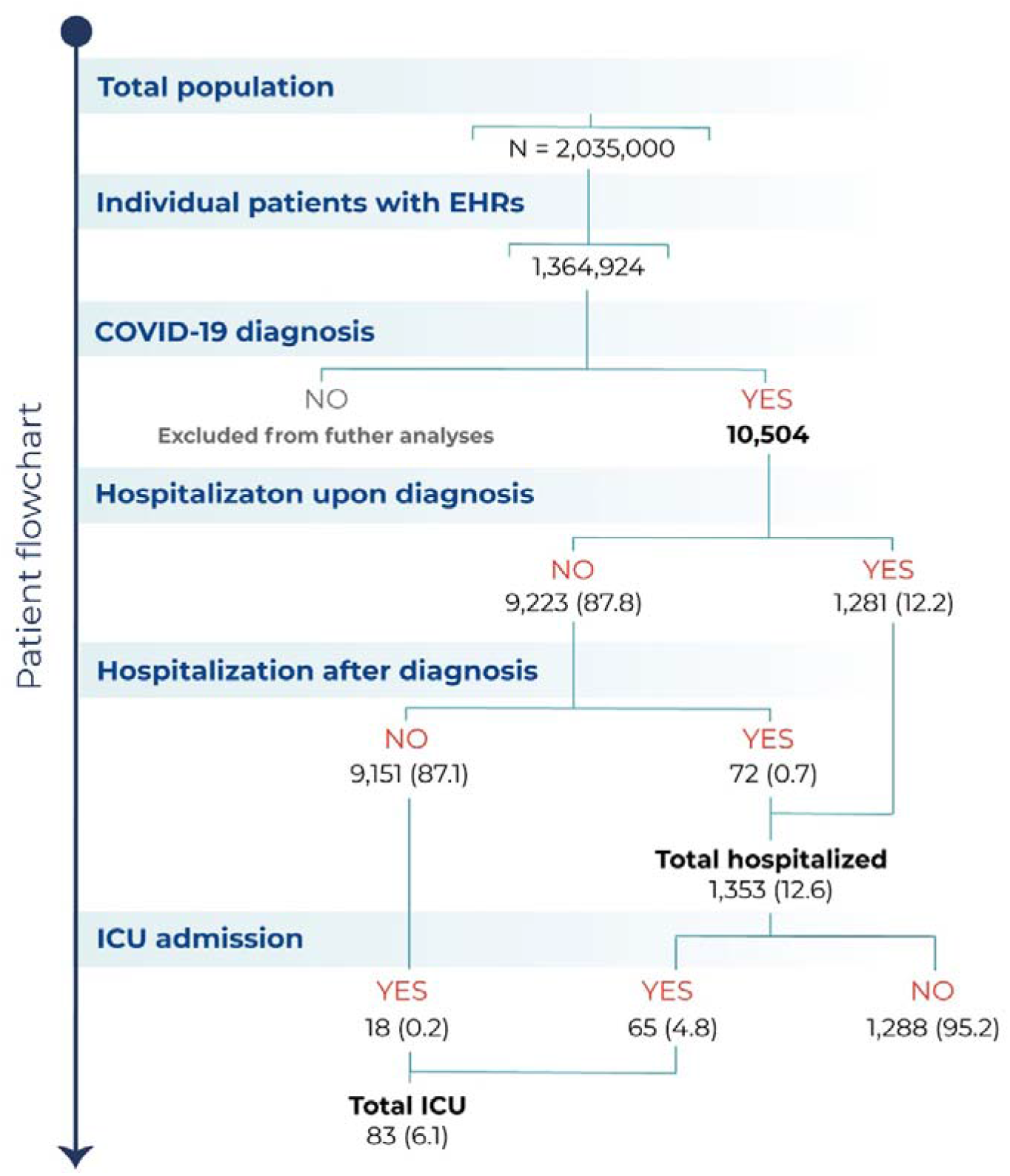
Patient flowchart. **Footnote**: Flowchart depicting the total number of inhabitants in the source population, the number (%) of patients with available EHRs analyzed, the number of patients diagnosed with COVID-19, and of those, the number of hospitalizations and ICU admissions.

COVID-19 patients were 52.5% males, with a mean±SD age of 58.2±19.7 years, (**Table 1**). Most COVID-19 patients were 50 years and older **(Figure 3)**. Upon diagnosis, the most common symptoms reported were cough, fever and dyspnoea (**Table 1**); notably, less than half of patients presented with these symptoms, probably due to the fact that most were attended in primary care. Further, respiratory crackles, myalgia, and diarrhoea were identified in 5% or more of cases, while other respiratory and non-respiratory signs and symptoms were less common. Sex-dependent differences in sign and symptom frequencies upon diagnosis are shown in **Table 1**. Of note, we observed subtle increases in frequency of diarrhoea, myalgia, headache, chest pain, and anosmia in female COVID-19 patients, while men showed significant increases in fever, dyspnoea, respiratory crackles, ronchus, lymphopenia, and tachypnoea (all p<0.05).

**Table 1.** Baseline demographics and clinical data upon diagnosis.

|  | Female<br>n = 4,984 | Male<br>n = 5,519 | TOTAL<br>n = 10,504 | p-<br>value* |
| --- | --- | --- | --- | --- |
| **Sex** |  |  |  |  |
| Female |  |  | 4,984(47.4) |  |
| Male |  |  | 5,519(52.5) |  |
| Unknown |  |  | 1(0.0) |  |
| **Age (in years)** |  |  |  |  |
| Mean(SD) | 57.4(20.0) | 59.0(19.5) | 58.2(19.7) | <0.001 |
| Median(Min-Max) | 58.0(0.0-100.0) | 60.0(0.0-102.0) | 59.0(0.0-102.0) |  |
| (Q1-Q3) | (44.0-73.0) | (46.0-74.0) | (45.0-73.0) |  |
| **Signs and Symptoms n(%)** |  |  |  |  |
| Cough | 2,482(49.8) | 2,760(50.0) | 5,243(49.9) | 0.8453 |
| Fever | 2,120(42.5) | 2,783(50.4) | 4,904(46.7) | <0.001 |
| Dyspnoea | 1,476(29.6) | 1,818(32.9) | 3,294(31.4) | <0.001 |
| Respiratory crackles | 849(17.0) | 1,085(19.7) | 1,934(18.4) | <0.001 |
| Diarrhoea | 556(11.2) | 543(9.8) | 1,099(10.5) | 0.03 |
| Myalgia | 467(9.4) | 451(8.2) | 919(8.7) | 0.0326 |
| Headache | 462(9.3) | 302(5.5) | 764(7.3) | <0.001 |
| Rhonchus | 279(5.6) | 414(7.5) | 693(6.6) | <0.001 |
| Chest pain | 287(5.8) | 267(4.8) | 554(5.3) | 0.039 |
| Lymphopenia | 196(3.9) | 346(6.3) | 542(5.2) | <0.001 |
| Wheezing | 194(3.9) | 195(3.5) | 389(3.7) | 0.3567 |
| Tachypnoea | 135(2.7) | 203(3.7) | 338(3.2) | 0.0059 |
| Anosmia | 166(3.3) | 134(2.4) | 300(2.9) | 0.0066 |
| Sore throat | 69(1.4) | 57(1.0) | 127(1.2) | 0.118 |
| Ageusia | 33(0.7) | 32(0.6) | 65(0.6) | 0.68 |
| Dysphagia | 19(0.4) | 28(0.5) | 47(0.4) | 0.4119 |
| Neuralgia | 19(0.4) | 22(0.4) | 41(0.4) | 1 |
| Splenomegaly | 8(0.2) | 14(0.3) | 22(0.2) | 0.4071 |
| Hepatomegaly | 2(0.0) | 6(0.1) | 8(0.1) | 0.3586 |
| **Comorbidities n(%)#** |  |  |  |  |
| Cardiovascular disease | 2,253(45.2) | 2,805(50.8) | 5,058(48.2) | <0.001 |
| Hypertension | 1,552(31.1) | 1,975(35.8) | 3,527(33.6) | <0.001 |
| Ischemic stroke | 91(1.8) | 163(3.0) | 254(2.4) | <0.001 |
| Heart Disease | 1100(22.1) | 1539(27.9) | 2639(25.1) | <0.001 |
| Ischemic heart disease | 152(3.0) | 475(8.6) | 627(6.0) | <0.001 |
| Heart failure | 243(4.9) | 309(5.6) | 552(5.3) | 0.1063 |
| Diabetes | 689(13.8) | 957(17.3) | 1646(15.7) | <0.001 |
| Obesity | 479(9.6) | 457(8.3) | 936(8.9) | 0.0185 |
| Renal dysfunction | 271(5.4) | 493(8.9) | 764(7.3) | <0.001 |
| CKD | 171(3.4) | 323(5.9) | 494(4.7) | <0.001 |
| Depression | 484(9.7) | 219(4.0) | 703(6.7) | <0.001 |
| Chronic respiratory disease | 242(4.9) | 646(11.7) | 888(8.5) | <0.001 |
| Asthma | 496(10.0) | 263(4.8) | 759(7.2) | <0.001 |
| COPD | 126(2.5) | 549(9.9) | 675(6.4) | <0.001 |
| Obstructive sleep apnea syndrome (OSA) | 69(1.4) | 143(2.6) | 212(2.0) | <0.001 |
| Bronchiectasis | 42(0.8) | 87(1.6) | 129(1.2) | <0.001 |
| Chronic Liver Disease | 36(0.7) | 75(1.4) | 111(1.1) | 0.002 |
| Cirrhosis | 16(0.3) | 35(0.6) | 51(0.5) | 0.0304 |
| HIV | 12(0.2) | 22(0.4) | 34(0.3) | 0.2113 |

**Footnote:** \*p-values from Yates-corrected $\chi^2$ test on percentage difference of female vs. male COVID-19 patients. All tests were performed individually for each variable (comorbidity or sign/symptom, where applicable). For numerical values (i.e., age), t-tests of difference between means were used. #List of medical conditions according to SNOMED CT terminology.

**Figure 3.**
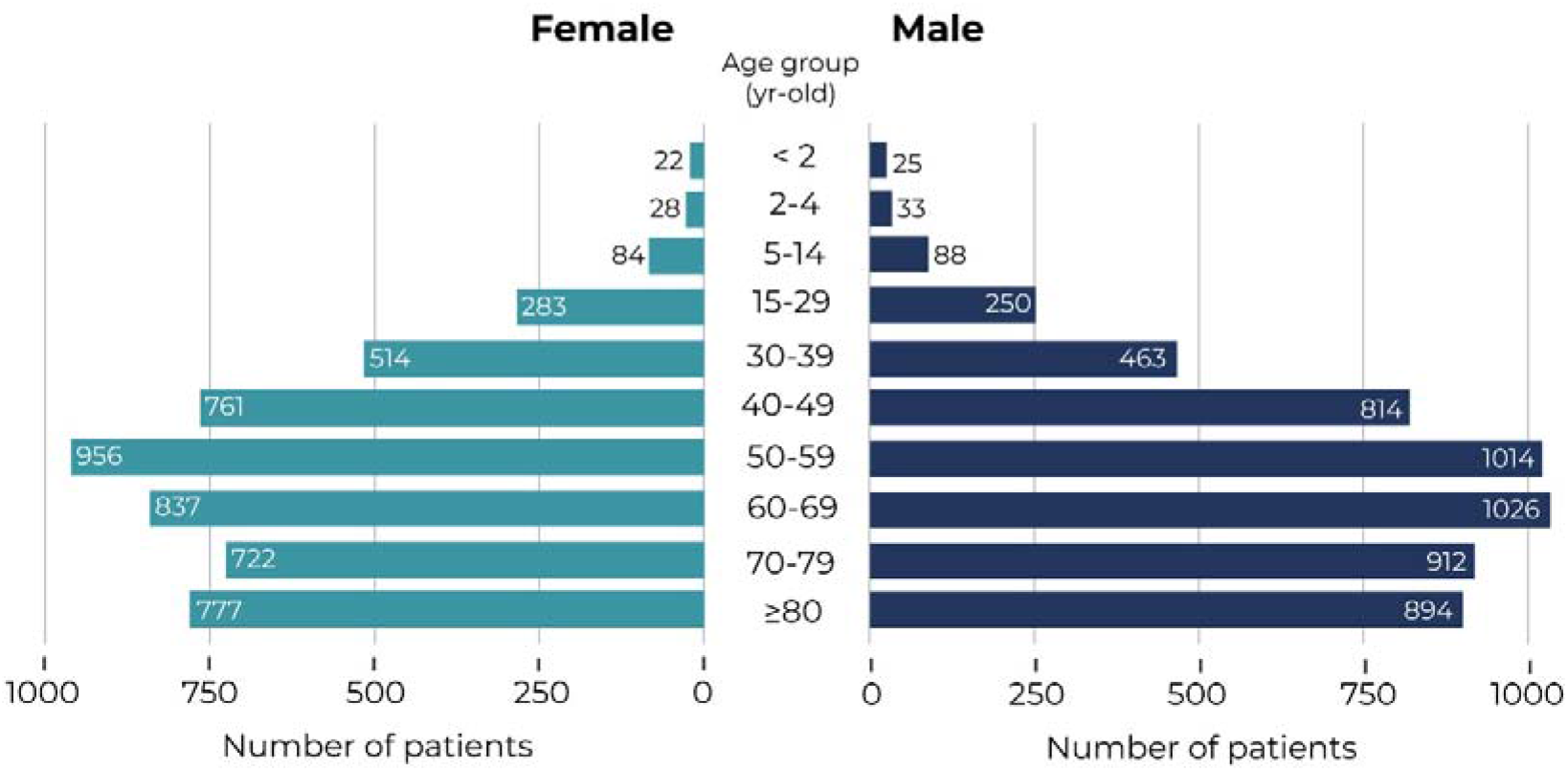
Age and Sex Distribution of COVID-19 patients. **Footnote**: Age distribution of incident cases of COVID-19 in females (left) and males (right) in the study population for the period comprised between Jan 1, 2020 and March 29, 2020.

Similarly, the most frequent comorbidities were cardiovascular disease (48.2% of patients) -mainly arterial hypertension (33.6%) and heart disease (25.1%)- and diabetes (15.7%) (**Table 1**). Regarding respiratory diseases, COPD was present in 6.4%, asthma in 7.2%, OSA in 2%, and bronchiectasis in 1.2% of patients. Sex-dependent differences in comorbidities upon diagnosis are also shown in **Table 1**; except for asthma, the frequency of all comorbidities was significantly higher in male than female COVID-19 patients (all p<0.05).

Next, we explored whether the distribution of comorbidities and sign/symptoms captured in the patients’ EHRs upon diagnosis differed between those COVID-19 patients who were admitted to the ICU vs. those who were not (**Table 2**). Regarding comorbidities, diabetes, obesity, cardiovascular disease (mainly hypertension), heart disease (mainly ischemic heart disease), and renal dysfunction were more common among those patients who were admitted to the ICU (all p < 0.01). As for signs and symptoms, cough, fever, dyspnoea, respiratory crackles, diarrhoea, tachypnoea, lymphopenia, and rhonchus were more frequent among ICU patients (all p < 0.05). Interestingly, respiratory diseases were not more frequent among patients who were admitted to the ICU (**Table 2**).

**Table 2.** Association between ICU admission and comorbidities/signs and symptoms upon diagnosis in patients with COVID-19.

| COMORBIDITIES |  |  |  | SIGNS AND SYMPTOMS |  |  |  |
| --- | --- | --- | --- | --- | --- | --- | --- |
| Condition<sup>#</sup> | No ICU<br>n(%) | ICU<br>n(%) | p-<br>value* | Sign or<br>Symptom | No ICU<br>n(%) | ICU<br>n(%) | p-<br>value* |
| Diabetes | 1613(15.5) | 33(39.8) | <0.001 | Cough | 5181(49.7) | 62(74.7) | <0.001 |
| Obesity | 917(8.8) | 19(22.9) | <0.001 | Fever | 4849(46.5) | 55(66.3) | <0.001 |
| Chronic<br>respiratory<br>disease | 883(8.5) | 5(6) | 0.548 | Dyspnoea | 3246(31.1) | 48(57.8) | <0.001 |
| COPD | 673(6.5) | 2(2.4) | 0.2029 | Respiratory<br>crackles | 1904(18.3) | 30(36.1) | <0.001 |
| Asthma | 750(7.2) | 9(10.8) | 0.2868 | Myalgia | 908(8.7) | 11(13.3) | 0.2066 |
| OSA | 211(2) | 1(1.2) | 0.8908 | Diarrhoea | 1084(10.4) | 15(18.1) | 0.0363 |
| Bronchiectasis | 129(1.2) | 0(0) | 0.6033 | Dysphagia | 47(0.5) | 0(0) | 1 |
| Cardiovascular<br>disease | 4998(48) | 60(72.3) | <0.001 | Wheezing | 383(3.7) | 6(7.2) | 0.1568 |
| Hypertension | 3487(33.5) | 40(48.2) | 0.0066 | Tachypnoea | 311(3) | 27(32.5) | <0.001 |
| Ischemic<br>stroke | 253(2.4) | 1(1.2) | 0.716 | Chest pain | 546(5.2) | 8(9.6) | 0.1237 |
| Heart Disease | 2604(25) | 35(42.2) | <0.001 | Lymphopenia | 524(5) | 18(21.7) | <0.001 |
| Ischemic<br>Heart<br>Disease | 616(5.9) | 11(13.3) | 0.0099 | Headache | 757(7.3) | 7(8.4) | 0.8442 |
| Heart failure | 548(5.3) | 4(4.8) | 1 | Rhonchus | 676(6.5) | 17(20.5) | <0.001 |
| Renal<br>dysfunction | 748(7.2) | 16(19.3) | <0.001 | Hepatomegaly | 8(0.1) | 0(0) | 1 |
| CKD | 488(4.7) | 6(7.2) | 0.4059 | Anosmia | 297(2.9) | 3(3.6) | 0.9317 |
| Chronic Liver<br>Disease | 109(1) | 2(2.4) | 0.502 | Ageusia | 65(0.6) | 0(0) | 0.9847 |
| Cirrhosis | 51(0.5) | 0(0) | 1 | Neuralgia | 41(0.4) | 0(0) | 1 |
| Depression | 699(6.7) | 4(4.8) | 0.6418 | Sore throat | 126(1.2) | 1(1.2) | 1 |
| HIV | 33(0.3) | 1(1.2) | 0.6536 | Splenomegaly | 21(0.2) | 1(1.2) | 0.4317 |

**Footnote:** \*p-values from Yates-corrected chi<sup>2</sup> test of difference between percentage of patients in either outcome group. All tests were performed individually for each variable (comorbidity or sign/symptom, where applicable). <sup>#</sup>List of medical conditions according to SNOMED CT terminology.

Finally, by using a machine-learning, data-driven algorithm, we identified that the combination of three easily available clinical variables, namely age, temperature, and respiratory frequency, was the most parsimonious predictor of ICU admission among COVID-19 patients (**Figure 4**). For this model, age and temperature were captured as continuous variables, whereas tachypnoea (yes/no) was defined as respiratory frequency of more than 20 breaths per minute. With accuracy, recall, and AUC values of 0.68, 0.71, and 0.76, respectively, the presented model reached optimal balance in terms of positive and negative predictive value for ICU admission. On the one hand, those younger than 56 years, without tachypnoea, and with temperature <39°C/102°F (entropy = 0, n = 145) (or >39°C/102°F without respiratory crackles), were free of ICU admission, (entropy = 0, n = 18). On the other hand, COVID-19 patients aged 40 to 70 years were likely to be admitted in the ICU if they presented with tachypnoea and delayed their visit to the ER after being seen in primary care (entropy = 0, n = 104). As stated in the Methods section, we performed an additional sensitivity analysis with different data sets to further validate the results of our predictive model. The independent data set of two provinces (Ciudad Real and Guadalajara, including a total of 753,408 individual patients, or 38% of the entire study sample from Castilla-La Mancha; **Figure 1** and **Supplemental Table S1**), was used to retrain our algorithm to identify ICU admission at onset; validation was performed in the remaining three provinces. As shown in **Supplemental Figure S1**, the new decision tree identified the same relevant clinical variables, that is age, tachypnea, temperature, and respiratory crackles/ronchus with similar (but not identical) thresholds in some of them. This additional model reached values of accuracy, recall, and AUC of 0.85, 0.57, and 0.84, respectively, thus providing additional proof of validity for our main findings.

**Figure 4.**
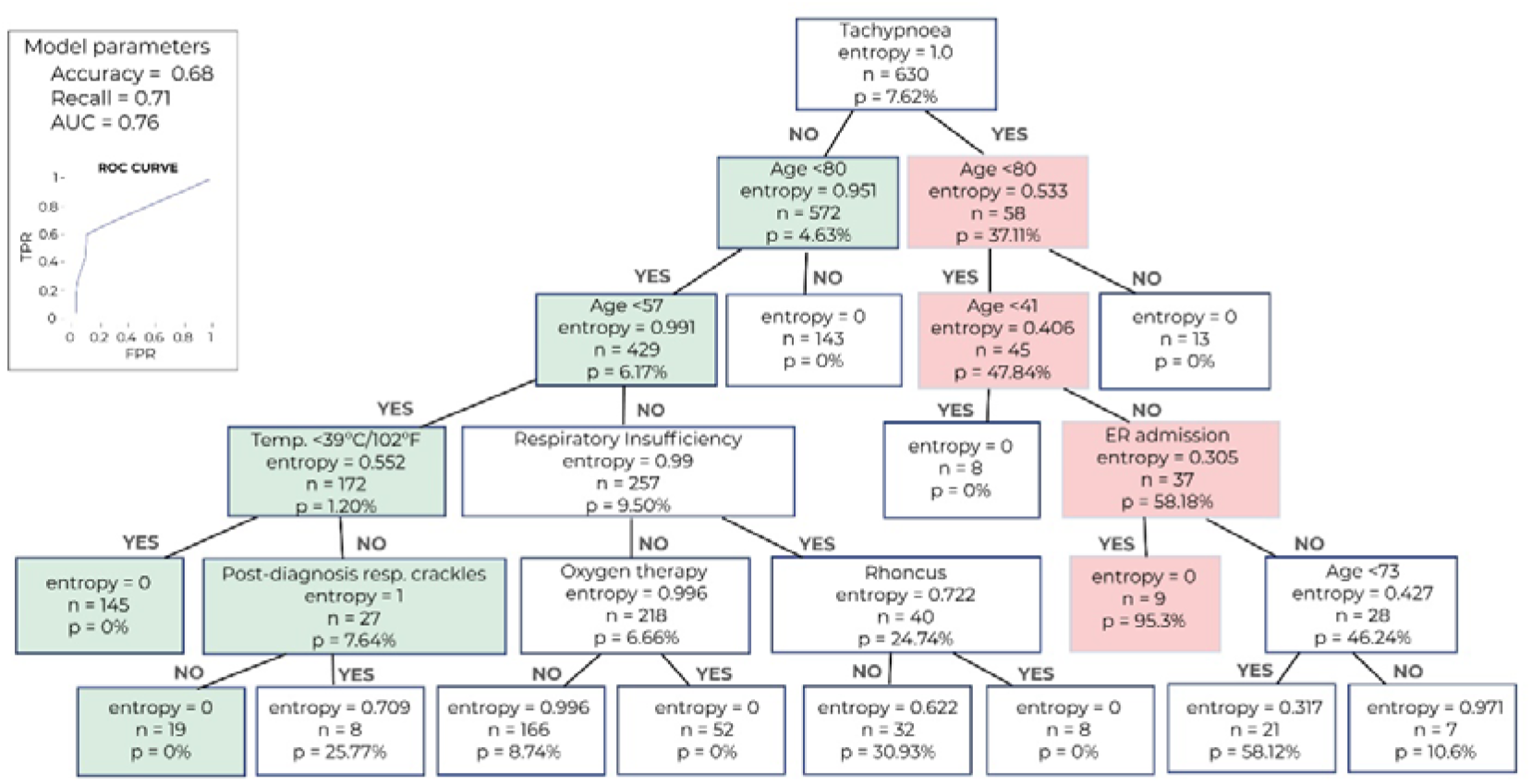
Decision tree of relevant clinical variables for the prediction of ICU admission in COVID-19 patients. **Footnote**The combination of three easily available clinical variables, namely age, temperature, and respiratory frequency, was the most parsimonious predictor of ICU admission among COVID-19 patients. The number of patients, probability (p) of ICU admission predicted by the model, and level of entropy (a measure indicating how mixed or pure the classification is, where 0 indicates optimal separation of classes) are indicated in each box. The green pathway indicates that those patients with no tachypnoea, younger than 56 years old, and with temperature less than 39°C/102°F (OR more than 39°C/102°F without respiratory crackles), did not require ICU admission. On the contrary, the red pathway indicates that patients aged 40-79 years, who presented with tachypnoea, and delayed their visit to the ER after being seen in primary care, were likely to be admitted in the ICU. For this model, we obtained accuracy, recall, and AUC values of 0.68, 0.71, and 0.76, respectively (top right panel). See text for further details.

## DISCUSSION

Recent technological advances allow for the optimal and rapid extraction, integration, and analysis of the unique and massive amount of untapped medical knowledge captured in the EHRs. This possibility is particularly meaningful when the clinical question at hand requires collecting data from a large number of patients in a very limited amount of time, as is the case with the newly described COVID-19 pandemic.

By anonymously accessing the clinical information of more than 10,000 anonymous patients with COVID-19 (a number that largely surpasses samples included in recent reports about the disease[27, 28]), we were able to describe their demographic and clinical characteristics, their clinical journey, and the statistical relationship between the most common symptoms and comorbidities on admission, and COVID-19 prognosis (i.e., ICU admission). There were subtle differences in clinical symptoms at onset by sex, while all comorbidities (but asthma) were significantly higher in male than female COVID-19 patients; these and other findings should be replicated in clinical series elsewhere.

The variables identified in our ICU admission model (i.e., age, temperature, and tachypnoea) are clinically relevant as they are readily available and easily observable in the everyday practice with COVID-19 patients. Although tachypnea is not an exclusive manifestation of COVID-19 and can be present in patients suffering from other respiratory diseases (i.e., pneumonia), our model suggests that this sign (in combination with age and temperature) is the most reliable predictor of ICU use over other common symptoms and signs such as cough, dyspnea, or respiratory crackles.

In addition, given that the stability and capacity of ICUs worldwide is threatened by the rapid spread of the disease, the identification of individual factors that predict ICU admission may not only improve patient management but also optimize healthcare resource use and planning.

Further applied to other national and international healthcare networks, the tools and methodology presented here can potentially characterize and predict the prognosis of COVID-19 in a timely and unprecedented manner. As recently pointed out[29, 30], there might be value in the application of artificial intelligence to the current COVID-19 pandemic, not only to predict outbreaks[31] or read chest CT scans[32], but also to disentangle COVID-19’s clinical onset and natural history in nearly real-time. Indeed, classical methods would have required months of questionnaire-based data collection and questionnaire validation, along with multiple Ethics Board approvals and other practical hurdles, all saved with our current approach.

In the race against COVID-19[33], where the goal is to curb the pandemic, it is imperative to leverage big data and intelligent analytics for the betterment of public health. However, it is of the utmost importance not to neglect privacy and public trust, to keep best practices, and to maintain responsible standards for data collection and data processing at a global scale[34].

### Strengths and Limitations

To our knowledge, this is the first study using NLP and machine learning to access real-world data in such a large COVID-19 population. Indeed, our state-ot-the-art methodology allowed for the rapid analysis of the unstructured free-text narratives in the EHRs of one million patients from the general population of the region of Castilla La-Mancha (Spain).

Our methodology combined modules for sentence segmentation, tokenization, text normalization, acronym disambiguation, negation detection, and a multi-dimensional ranking scheme; the latter involved linguistic knowledge, statistical evidence, and continuous vector representations of words and documents learned via shallow neural networks. When applied to EHRs, NLP enables *a)* access to entire track records for *all* patients in the target population, and *b)* the implementation of exploratory analysis to unravel associations between variables that have remained undetected with traditional research methods. By considering all possible patients with the target disease, the information and analyses used here (i.e., RWD and free-scale statistics) remained unbiased by the research question or the observers. Unlike classical statistical methods (e.g., logistic regression), the main advantage associated with the use of ML in this context is that it allows for the automatic detection of meaningful relationships between variables. For instance, if a given symptom (i.e., fever) is only relevant for certain patients (i.e., older than 50), techniques such as the classification trees used here are suitable to uncover this relationship. In this context, although the total number of patients that required ICU use in the training set was somewhat low (48 patients), the number of variables considered in the model was also very limited. In addition, the inclusion of a validation stage reduces the likelihood of overfitting. Ultimately, the use of classifications trees in this study (as opposed to other models such as Artificial Neural Networks) is especially appropriate in the clinical context because they are easily interpretable.

Regarding the geographical location of our participating hospital sites, it is worth mentioning that with a total of 1,364,924 patients from the region of Castilla La-Mancha (SESCAM Healthcare Network), our sample is representative of the Spanish population; Spain has been among the hardest hit countries by the pandemic, in terms of both total cases and mortality rates [35, 36], and this region in particular is the third most affected in the country, just behind Madrid and Catalonia. For this reason, we anticipate that the clinical conclusions drawn here are relevant for clinicians worldwide. Of note, ICU capacity in the region during the study period was not compromised yet, which protects against possible bias in our training data (all patients requiring intensive care were indeed admitted to the ICU).

The results and conclusions of the present study should be interpreted in light of the following limitations. First, we did not distinguish COVID-19 cases confirmed by laboratory results (i.e, RT-PCR) from those exclusively diagnosed through clinical observation (i.e., symptomatology, imaging and laboratory results). However, it should be noted that PCR and other rapid laboratory tests for the detection of SARS-CoV-2 were not routinely administered in Spain during the study period. In addition, this decision is supported by recent discussions on the clinical validity and relatively high sensitivity of symptom- and imaging-based identification of COVID-19 patients, especially in early stages of the disease[17, 19, 20]. Second, independent replications by different research groups in larger patient sets are needed to further support the clinical validity of our results.

Finally, future reports from the BIGCOVIData study may incorporate laboratory results and treatments, and contextualize the results presented here in a larger clinical picture[25].

We conclude that, in the largest series of COVID-19 patients attended during the first three months of the pandemic in Spain, 6% of all hospitalized patients required ICU; and that a combination of easily obtained clinical variables, namely age, fever, and tachypnoea predicts which COVID-19 patients require ICU admission.

## Data Availability

Tabulated data is available upon request to the authors and subject to all EU regulations

https://bigcovidata.savanamed.com/

## Acknowledgments

We thank all the Savaners for helping accelerate health science with their daily work. This would have not been possible without every single team member. We also thank SESCAM (Healthcare Network in Castilla-La Mancha, Spain) for its participation in the study and for supporting the development of cutting-edge technology in real time.

